# Differences in metatarsal structure and mechanical behavior are small in runners with and without acute metatarsal bone stress injury

**DOI:** 10.1101/2025.03.04.25323333

**Authors:** Andrew R. Wilzman, Bryhannah A. Young, Irene S. Davis, Adam S. Tenforde, Karen L. Troy

## Abstract

**Purpose:** To investigate differences in metatarsal bone structure and training habits in runners with and without a recent metatarsal bone stress injury (BSI).

**Methods:** Fifty-four runners (14 male/40 female, age 25.8±7.3 yrs) who ran 47±32 kilometers weekly participated in this study. Training and injury history data were collected, along with CT images from metatarsals 2-4 of the non-injured foot of recently injured runners (n=11, 5 male), and the left foot from the healthy runners (n=43, 9 male). Quantitative CT analysis was performed and subject-specific finite element (FE) models simulated a “virtual mechanical test” on each bone at a range of biomechanically relevant angles. Key FE outcomes included principal strains and a measure of total damaged volume, which is related to fatigue life.

**Results:** Injured runners reported significantly higher training volume (78.9±33.9 km/week) than healthy runners (39.2±20.2 km/week) and had lower BMI (21.3±1.7 vs. 22.7±2.6 kg/m2) but the groups were otherwise similar. In the female group, injured runners had significantly larger bone volume and BMC, similar bone strains, and significantly higher damaged volume metrics than healthy females. The FE simulations showed that decreasing the loading angle of the metatarsals by 10 degrees was associated with a 22% decrease in strain and damaged volume.

**Conclusion:** The metatarsals of injured and healthy runners are only slightly different from each other, and there are no obvious structural deficits in the injured runners. Other factors including training volume, footstrike biomechanics, and sex differences may explain BSI in this cohort. Interventions that decrease metatarsal loading angle or magnitude may reduce BSI risk by reducing bone microdamage.

## Introduction

Bone stress injuries (BSI) are overuse injuries caused by the repeated loading of bone during weight-bearing activities such as running, jogging, and marching [1]. Repetitive loads cause the accumulation of fatigue damage, which stimulates targeted remodeling. BSI occurs when the rate of damage exceeds the rate bone repair. Military professionals and athletes are at a higher risk for BSIs, with women being more susceptible to these injuries than men [2]. The metatarsals are amongst the most common sites of injury [1,3,4]. The foot was reported as the most common location in college athletes [5] and a separate report noted that 38% of all BSI sustained were localized to the metatarsals [6]. In high school athletes, over 18% of reported BSIs were recurrent, with metatarsals having >10% recurrence rate [7]. In the military, metatarsal BSIs represent the majority (58%) of total BSIs. Metatarsal BSI occurs throughout elite military training, suggesting that cumulative loading exposure contributes to this form of injury [8].

There are several known risk factors for metatarsal BSI, which can be broadly categorized as being biological/physiologic, training-related, and biomechanical in nature. Biological and physiologic risk factors include genetics, circulating hormone levels that affect bone metabolism, and dietary factors[9]. These affect the observed bone structure, as well as the ability of bone to repair damage. Training related factors are related to training volume, type, and habituation to changes in training. For example, the total number of miles run is strongly related to the development of BSI, with increased risk in those running over 64 km per week [10]. Rapid increases in training volume or the introduction of new activities can also lead to injury because the musculoskeletal system takes time to adapt. Biomechanical factors include footstrike pattern, foot posture (arch height/structure), and bone structure [11]. Collectively, demands to bone dictate the forces and moments applied to the metatarsals, as well as the resulting strains within the bone. Ultimately, it is the repeated cycles of high strain that cause BSI.

Because BSIs result from cumulative damage, BSI risk could potentially be explained by estimating metatarsal fatigue life. In general, the fatigue life of a material can be expressed as a function of peak-to-peak stress or strain and the number of cycles to failure [12]. In small beam specimens, bone fatigue failure life also depends on the volume of the specimen being tested, with larger specimens failing in fewer cycles than smaller specimens for a given stress/strain [13]. This consistent observation can be conceptualized as a “weakest link” phenomenon, where larger specimens have more opportunities for failure to occur than smaller specimens. This phenomenon has led to a well-supported theory that the total volume of highly stressed (or strained) material is a key determinant of bone fatigue life[13]. For example, Gargac et al. showed that the location of regions of cortical damage in rat femora subjected to fatigue bending loads could be best explained by the (local) contiguous volume of elements experiencing high (>0.5%, or 5000 με) strain [14]. Other studies have shown that strain magnitudes greater than 4,000 με cause bone damage [15] and that the total volume of highly strained bone can predict whole-bone fatigue life in rabbit tibiae [16]. Thus, strained volume may be a useful surrogate measure for estimating the fatigue life of bone during *in vivo* loading scenarios. Since bone material is anisotropic, it is important to consider the multiaxial stress state rather than a simple measure of maximum or minimum principal strains. Other studies have shown that the Tsai-Wu failure criterion, which accounts for bone yield in all three dimensions and in both tension and compression, can accurately model bone fractures during monotonic loading [17]. This concept could be expanded to examine the total amount of “damaged volume” in a bone as a surrogate measure for fatigue life.

While biomechanical factors have been described to contribute to metatarsal BSI, limited work has applied these principles to modeling risk for this form of injury in runners. Here, we sought to understand the importance of bone structure and its relation to training volume in the context of metatarsal BSI. We asked two questions about runners with and without a recent metatarsal BSI: (1) How does metatarsal bone structure relate to body size and training volume, and is this relationship similar in injured and non-injured (healthy) runners? And (2) Do injured runners have weaker metatarsals than healthy runners? We hypothesized that metatarsal structure would scale with body size (larger bones in bigger runners) but not training volume, and that this scaling would not be affected by injury status. We also hypothesized that injured runners would exhibit lower metatarsal bone quality and mechanical properties compared to healthy runners.

## Methods

### Participant Overview

Fifty-four runners (14 male, 40 female, age: 25.8±7.3 years-old, height: 170±9 cm, mass: 64.8±9.8 kg, BMI: 22.5±2.7 kg/m^2^) were analyzed in this study. Participants reported running an average of 47±28 kilometers weekly over the last 12 months. All participants provided written informed consent for this institutionally approved study. Runners with no prior history of BSI were considered “healthy”. Runners with a unilateral metatarsal bone stress injury that was diagnosed within the past month were considered “injured”. Here, we analyzed the non-injured feet of 11 injured runners with unilateral injuries (6 female, 5 male), and the left feet from 43 healthy runners (34 female, 9 male), to achieve a dataset consisting of 54 feet. This sample was determined based on an *a priori* power analysis to detect an effect size of 0.9, which intentionally included more healthy than injured runners to increase statistical power. We focused recruitment on females because they sustain BSI at 2-3 times the rate of males [2] All participants completed enrollment forms documenting health metrics, [18]running history, recent running activity/statistics and information of current injury (if applicable).

### Imaging

The metatarsals were imaged with computed tomography. All images were acquired using an Xtreme CT I (Scanco Medical, Switzerland) with acquisition settings of 59.4 kV and 900 mA, 0.246 mm voxel size (Figure 1). A total scan length of 110 mm was acquired to encompass all five metatarsals and resulted in an effective radiation dose of 6 millirem. Images were checked for quality and motion artifact and removed if there was obvious blurring or misalignment between sequential slices that could not be corrected with image registration. Metatarsals two, three, and four (MT2, MT3, MT4) were individually segmented from the original whole foot scans and imported into Materialise Mimics software for QCT analysis and finite element model generation (Materialise v23.0, Leuven, Belgium). These metatarsals were selected based on being the most frequently injured in runners [1,3].

**Figure 1.**
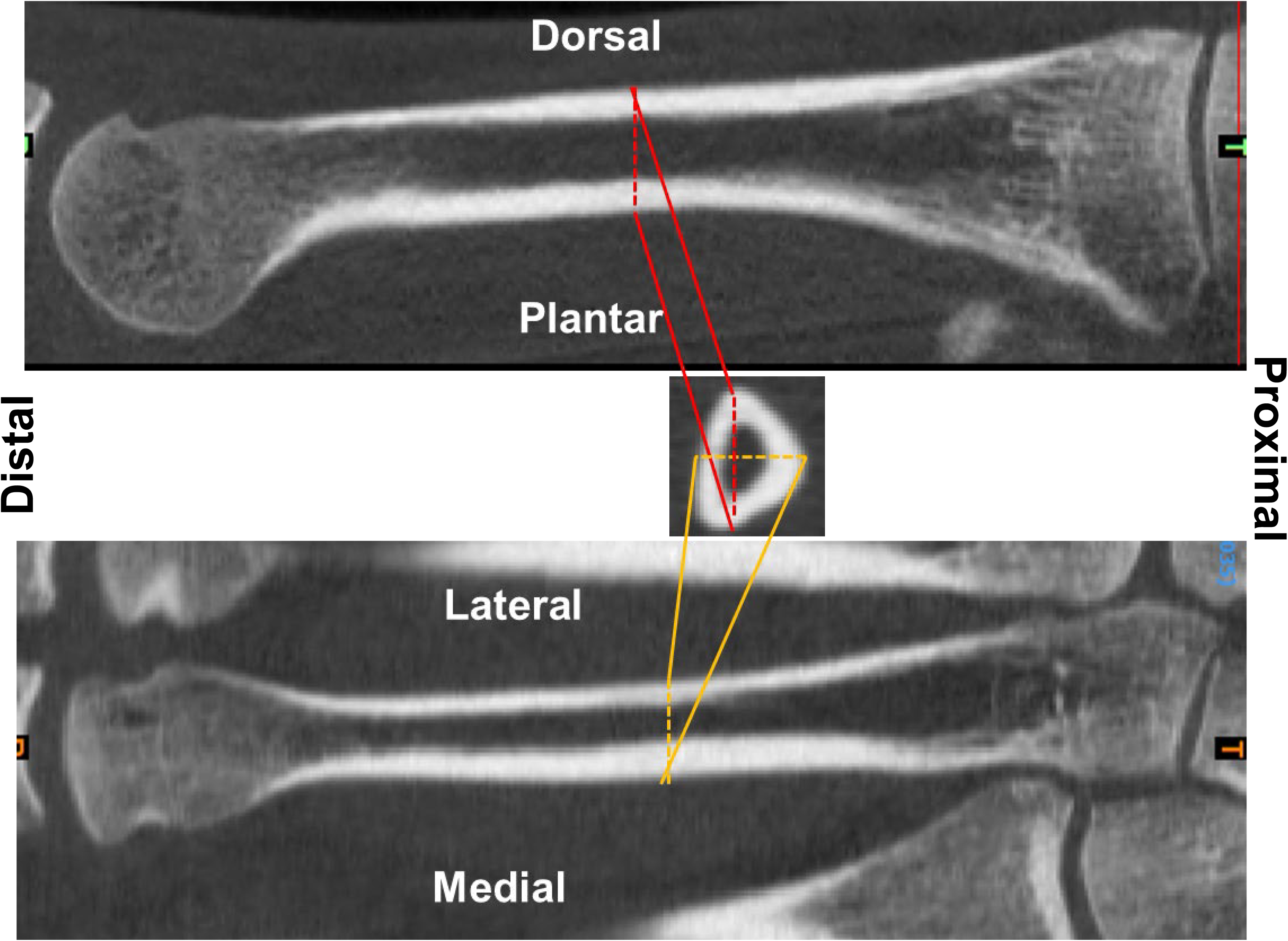
Cross-sectional views for a typical second metatarsal analyzed in this study. Images were acquired with a Scanco XtremeCT I (voxel size: 246 mm) 60 kVp 900 mA.

### QCT Analysis

Segmented metatarsal masks were exported from Mimics (v26.0, Materalise, UK) into custom Matlab code for QCT analysis. Based on our previously established methods [18,19], we calculated the following parameters from each entire metatarsal: integral bone mineral content (BMC; g), bone volume (BV; cm3), bone mineral density (BMD; g/cm3), average cross sectional area (CSA; mm2), compressive strength index (CSI; g^2^/mm^4^), bending strength index (BSI; cm3), and the average mass-weighted maximum (I_max_; mg*cm^2^), minimum (I_min_; mg*cm^2^) and polar moments of inertia throughout the length of the bone (J_0_; mg*cm^2^). To further explore differences along the length of the metatarsals, we calculated CSI, inertias, and the ratio of I_max_/I_min_ as a function of percent metatarsal length (from proximal to distal).

### Finite Element (FE) Modeling

Each metatarsal was processed through 3-Matic (v18.0, Materialise, UK) to generate size-adaptive, 10-node tetrahedral elements with a maximum edge length of 2 mm. Linear-elastic material properties were assigned based on a metatarsal-specific density-modulus optimization method proposed by Fung [20] (Equations 1-3), along with previously established orthotropic relationships [20,21].

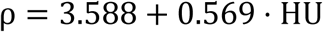

Equation 1. Density (ρ) is established as a linear relationship with Hounsfield Units (HU) using a calibration phantom.

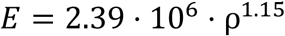

Equation 2. Compressive elastic modulus (E) is defined as an exponential function of density

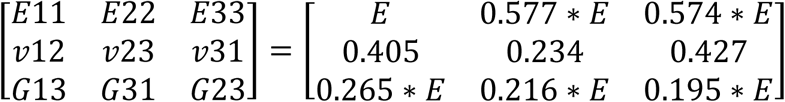

Equation 3. Orthotropic material properties are defined as a function of longitudinal compressive elastic modulus (E11) [20,21].

A series of simulations were run to quantify metatarsal mechanical behavior over a range of biomechanically relevant loading angles. To accomplish this, elements within 20 mm of the proximal end of each metatarsal were fully constrained. A force was applied at varying angles (70°, 65°, 60°, 55°, and 50°) to the most distal node, representing typical distal metatarsal loading vector directions that occur during the stance phase of gait due to the combined actions of the ground reaction force, intrinsic toe flexors, and plantar fascia [22]. Nodes located within a 5 mm radius of the force location were kinematically coupled (Figure 2). Each simulation consisted of a virtual mechanical test in which the magnitude of the applied force was arbitrarily assigned as 17.5% of each participant’s body weight, because this falls within the range of estimated metatarsal reaction forces during walking [22].

**Figure 2.**
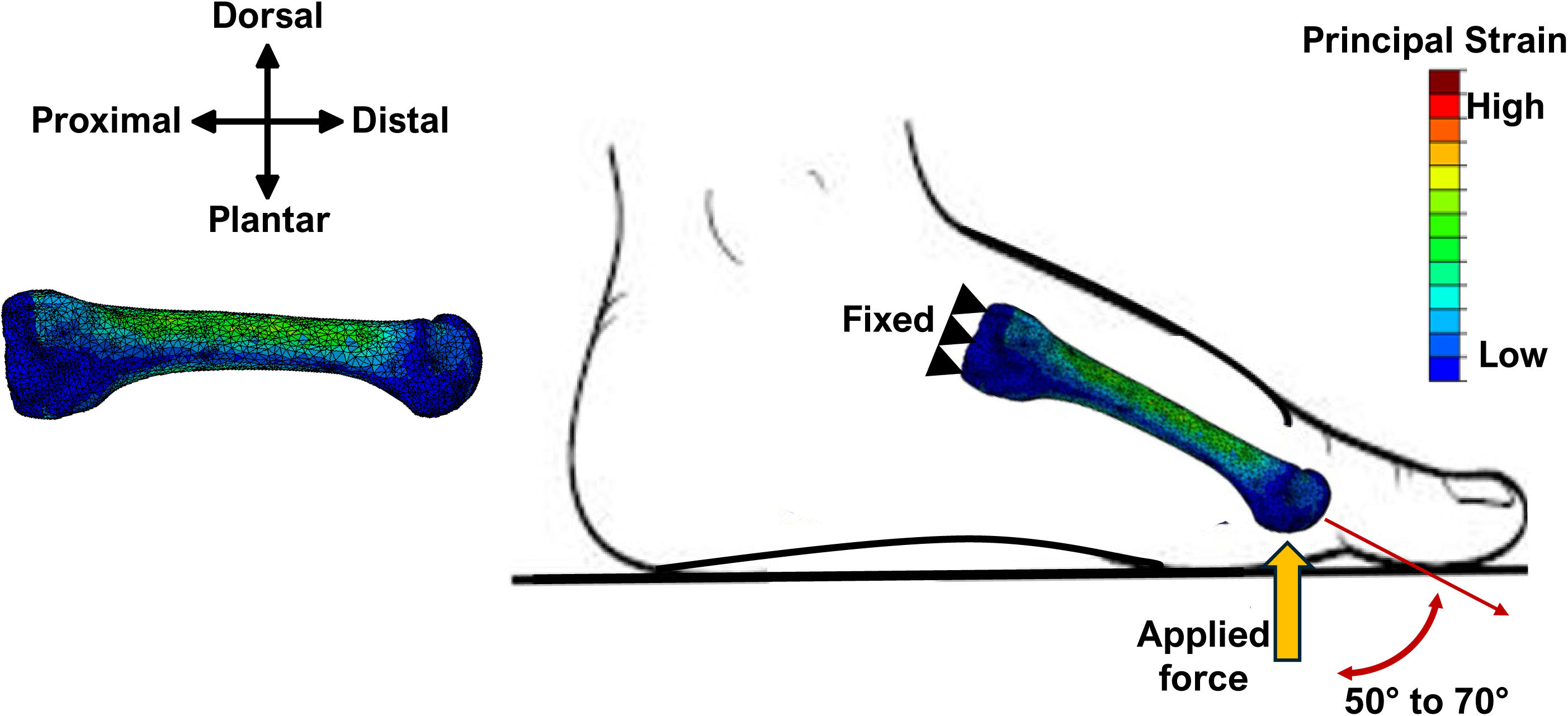
FE modeling setup used to stimulate the various loading angles. The yellow arrow represents the resultant force applied on the plantar surface on the distal end.

### FE Simulation Outcomes

A total of 810 simulations were run. For each, maximum and minimum principal strains of all elements five or more millimeters away from constrained nodes were recorded, to eliminate artificially high strains resulting from edge effects at the boundaries. Effectively, this yielded outcomes for elements within the diaphysis and distal end of the bone.

The multidirectional strain state of each element was then compared to the Tsai-Wu failure criterion with limits measured in human cortical bone [17,23–25]. Since the model used inhomogeneous material properties, a strain-based version of Tsai-Wu was calculated, which allowed for scaling of failure stresses in proportion to the individual element modulus values. Any element whose multiaxial strain state resulted in a criterion value greater than 1 was considered damaged. For each simulation we calculated the total damaged volume, which was the sum of the volumes of all damaged elements.

### Statistical Analysis

We first calculated descriptive statistics for all variables and performed a 2-way ANOVA to examine differences in demographics (age, height, mass, BMI, weekly distance) between injury groups and sexes. Next, we calculated Pearson correlations between all QCT variables and body size (height, mass) or weekly distance. To test for differences in QCT outcomes between injured/healthy runners and sexes, we used mixed linear models. Metatarsal number was considered a repeated factor, sex and prior BSI status were between-participant factors, and height and mass were included as covariates for all outcomes that were significantly correlated with body size. We also examined female participants only, given the large portion of women who participated in this study (n=40). To further explore whether specific regions of each metatarsal differed between the two groups, we used statistical parametric mapping (SPM) with a t-test to compare density-weighted moments of inertia and the ratio of I_max_/I_min_ as a function of bone length. A similar mixed linear model approach was used to examine FE outcomes but we treated each metatarsal separately so that loading angle could be a repeated measure. Additionally, since the boundary conditions applied to the FE models were already scaled for body size, we did not include height or weight as covariates in these models.

## Results

### Characteristics of Injured vs. Healthy Runners

Injured and healthy runners were similar in age, height, weight. However, injured runners had lower BMI and higher weekly training distances than healthy runners (p<0.05, Table 1). The injured runners had injuries on MT2 (n=5), MT3 (n=5) and MT4 (n=1).

**Table 1.** Demographics and weekly training distance for all study participants.

|  | Injured | n | age<br>(years) | height (cm) | mass (kg) | BMI (kg/m<sup>2</sup>) | weekly distance<br>(km) |
| --- | --- | --- | --- | --- | --- | --- | --- |
| **Female** | **No** | 34 | 27.0 (8.1) | 164.8 (5.4) | 61.4 (7.8) | 22.5 (2.5) | 42.9 (20.9) |
|  | **Yes** | 6 | 24.5 (7.0) | 172.0 (9.1) | 62.0 (4.2) | 21.0 (2.1) | 78.0 (35.2) |
| **Male** | **No** | 9 | 21.6 (2.0) | 180.0 (9.1) | 76.6 (11.2) | 23.6 (3.0) | 48.3 (26.4) |
|  | **Yes** | 5 | 24.2 (3.6) | 181.5 (5.6) | 70.9 (5.5) | 21.5 (1.0) | 80.0 (36.2) |
| **All** | **No** | 43 | 25.8 (7.6) | 168.1 (8.9)* | 64.7 (10.6)* | 22.7 (2.6)<sup>\$</sup> | 39.2 (20.2)<sup>\$</sup> |
| | **Yes** | 11 | 23.9 (5.5) | 176.3 (7.1)* | 66.0 (6.5)* | 21.3 (1.7)<sup>\$</sup> | 78.9 (33.9)<sup>\$</sup> |

\* p<0.001 for Female vs. Male
<sup>\$</sup> p<0.05 for healthy vs. injured

### Relationships between body size, weekly distance, and QCT outcomes

All QCT outcomes were related to body height and mass (p<0.001), with the exception of BMD (Figure 3). These relationships were not affected by injury status. Weekly training distance showed no relationship to any of the QCT outcomes.

**Figure 3.**
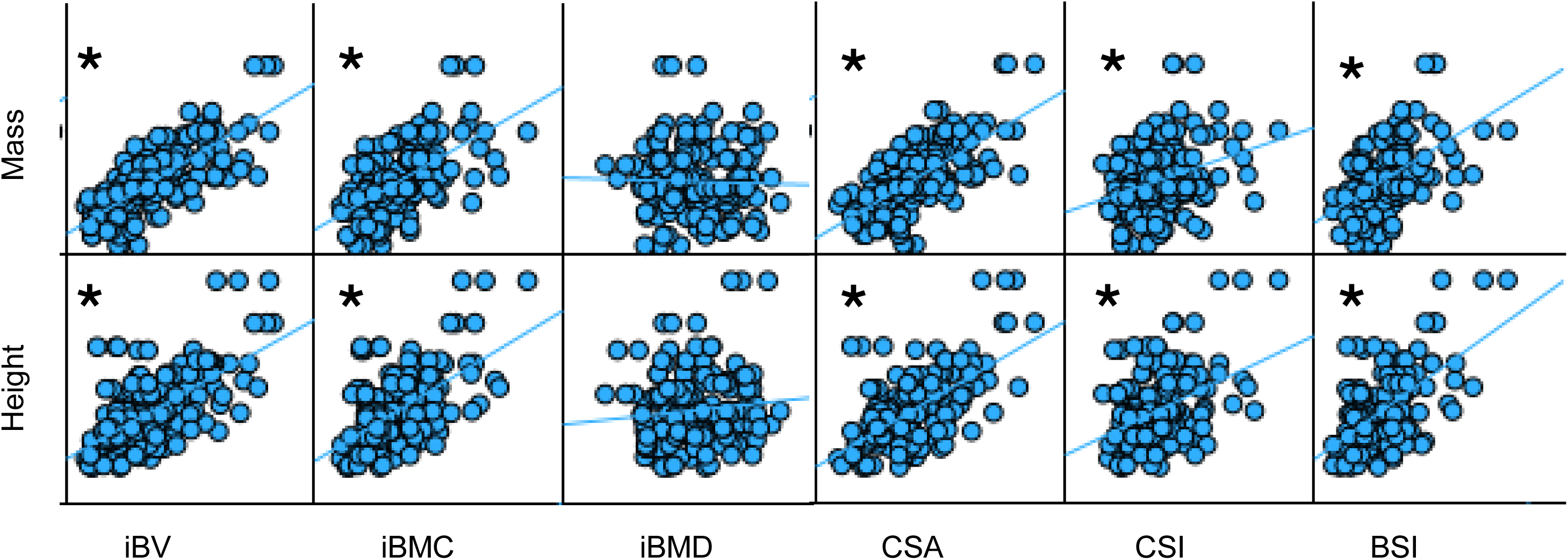
Scatter plots showing the relationships between body height and mass versus the QCT parameters. Significant relationships are indicated with a *.

### Bone Structure: QCT outcomes in Male and Female Injured vs. Healthy Runners

All bone size and strength indicators were different between male and female runners (p<0.001), with significant interactions between sex and injury status (p≤0.006 except for CSI, Figure 4). In general, injured female runners had greater bone volume, CSA, and strength indices compared to healthy female runners, while male runners showed the opposite trend. BMD was not different between sexes or injured/healthy groups. Furthermore, each of the metatarsals were different from each other (p<0.001), with interactions by metatarsal and injury status for I_max_ and I_max_/I_min_ variables. *Post hoc* analysis with Bonferroni corrections indicated that the differences were largely attributed to group differences in MT4. The SPM analysis revealed differences in the mid-shaft geometry of MT4 between injured and healthy runners. Specifically, injured runners had greater I_max_ and I_max_/I_min_ ratios from around 30-70% of the bone length (Figure 5).

**Figure 4.**
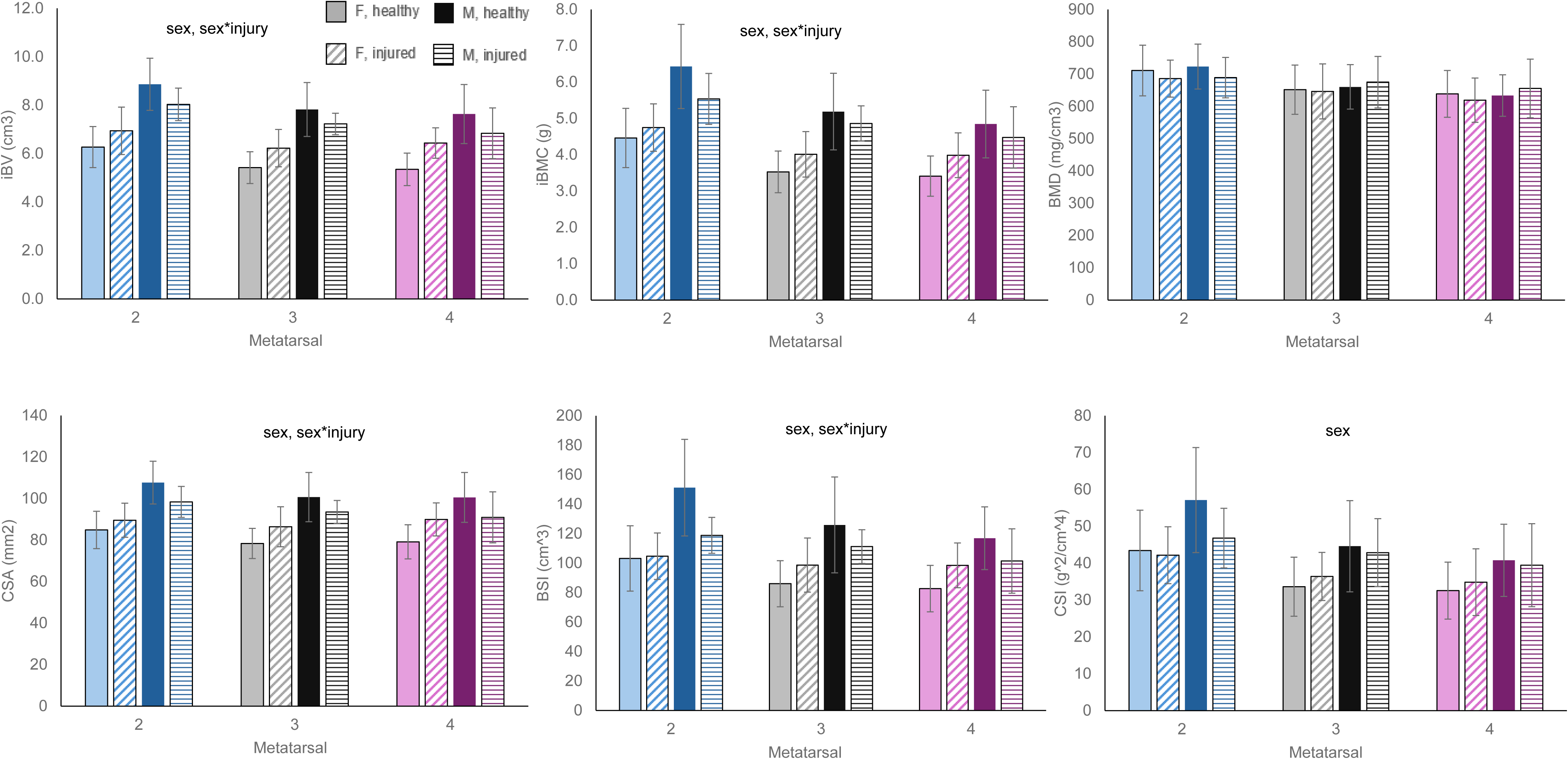
QCT parameters for each metatarsal in female and male healthy and injured participants. MT2 (blue), MT3 (black) and MT4 (purple) were all significantly different from each other. iBV, iBMC, CSA and BSI all showed significant differences between the sexes, as well as significant sex*injury status interactions. CSI only showed differences between the sexes. Error bars indicate SD.

**Figure 5.**
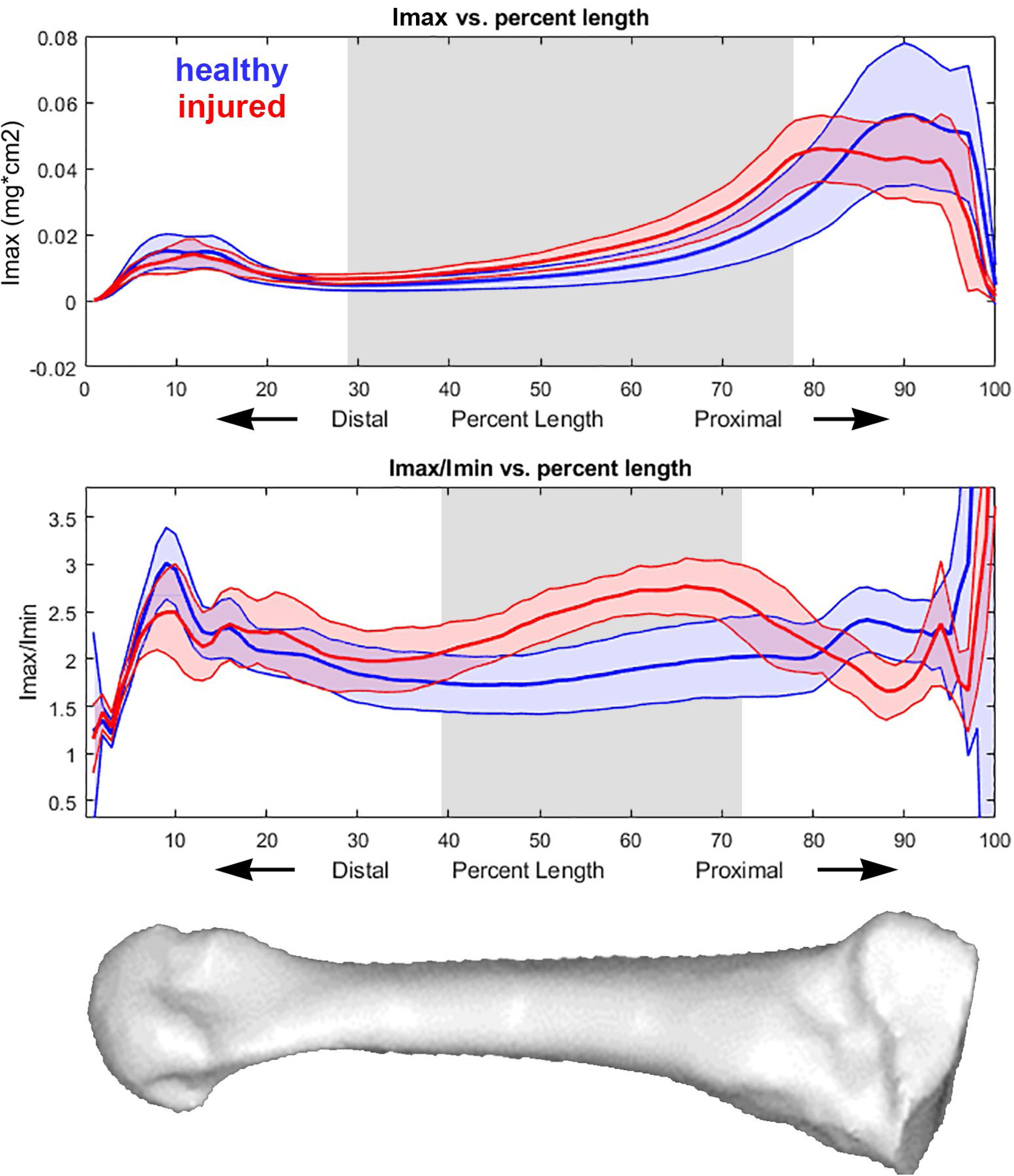
(Top) I_max_ as a function of percent bone length, and (middle) I_max_/I_min_ as a function of percent bone length. The bottom image is provided to orient the reader to where along the metatarsal bone each location corresponds. Red shows injured participants and blue shows healthy participants, with shaded regions are mean ± SD. The vertical gray shaded regions indicate those areas for which there are significant differences between injured and healthy participants.

### Bone mechanical behavior: distribution of strain and damaged volume

Due to the canteliever boundary conditions simulated in the virtual mechanical test, the greatest tensile loads occurred at the plantar surface while the greatest compressive loads occurred at the dorsal surface of each metatarsal. Damaged elements were located at the high-strain cortical surface locations, primarily towards the proximal end (Figure 6). The 17.5% bodyweight force produced peak strain values ranging from 6000 to 10,000 micro-strain in both tension (positive values) and compression (negative values).

**Figure 6.**
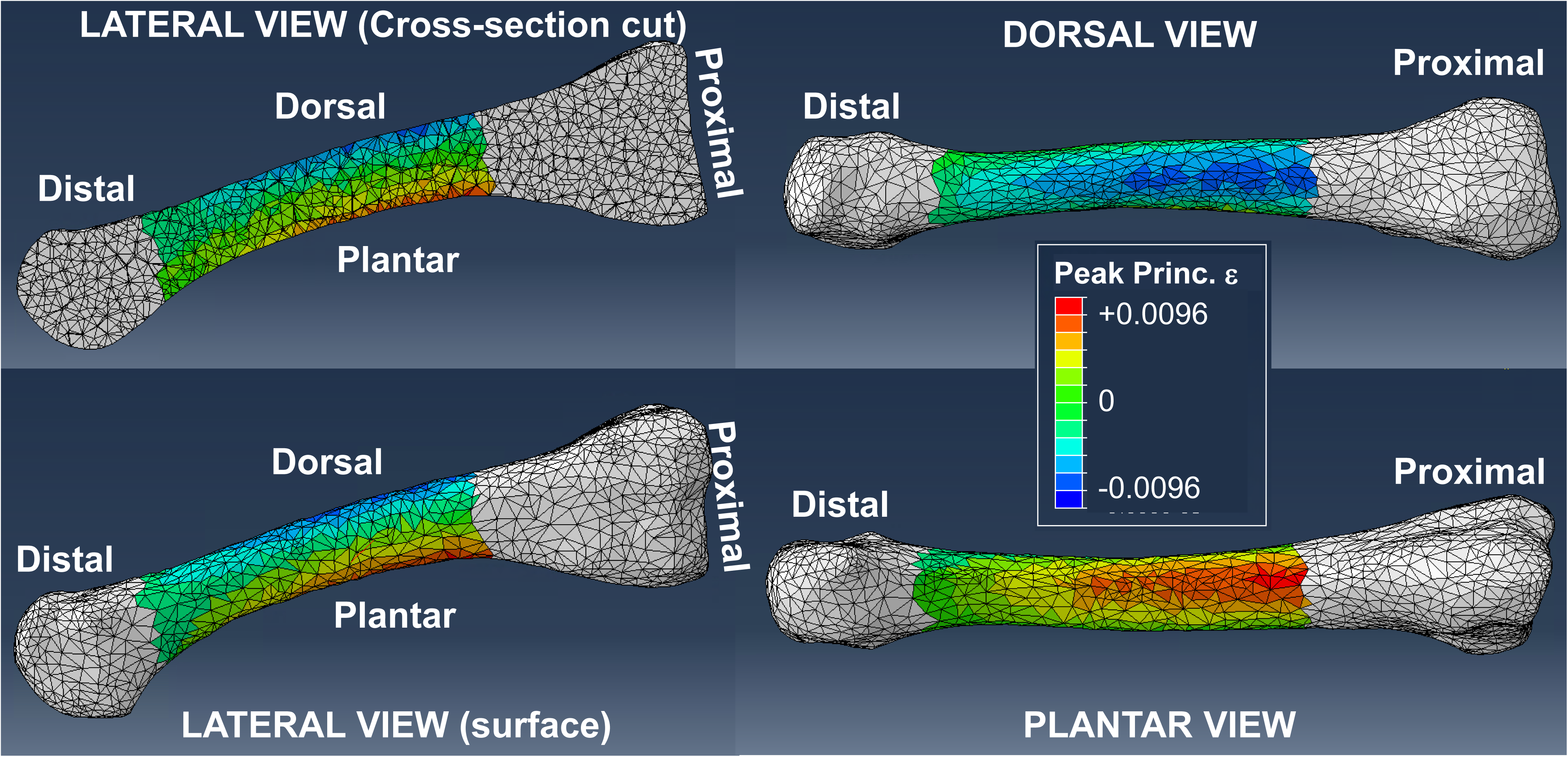
Typical FE model results showing the strain distribution within and on the surface of a third metatarsal. The dorsal surface experienced compression (indicated with negative strain values) while the plantar surface experienced tension (positive strain values). To avoid artifact from the boundary conditions, the elements shown in white are not analyzed in this study.

### Bone Strength: Metatarsal Strains in Male and Female Injured vs. Healthy Runners

Maximum and minimum principal strains and damaged volume all increased in association with higher loading angle (p<0.001). This was particularly apparent with damaged volume, which consistently increased with higher loading angle (Figure 7). Damaged volume was also different between male and female participants, with damaged volume being consistently larger in males (p<0.001). When only female particpants were analyzed, we observed that damaged volume was higher in the injured participant MT2 and MT4, especially at higher loading angles. Principal strains were similar between sexes and injury groups across all metatarsals.

**Figure 7.**
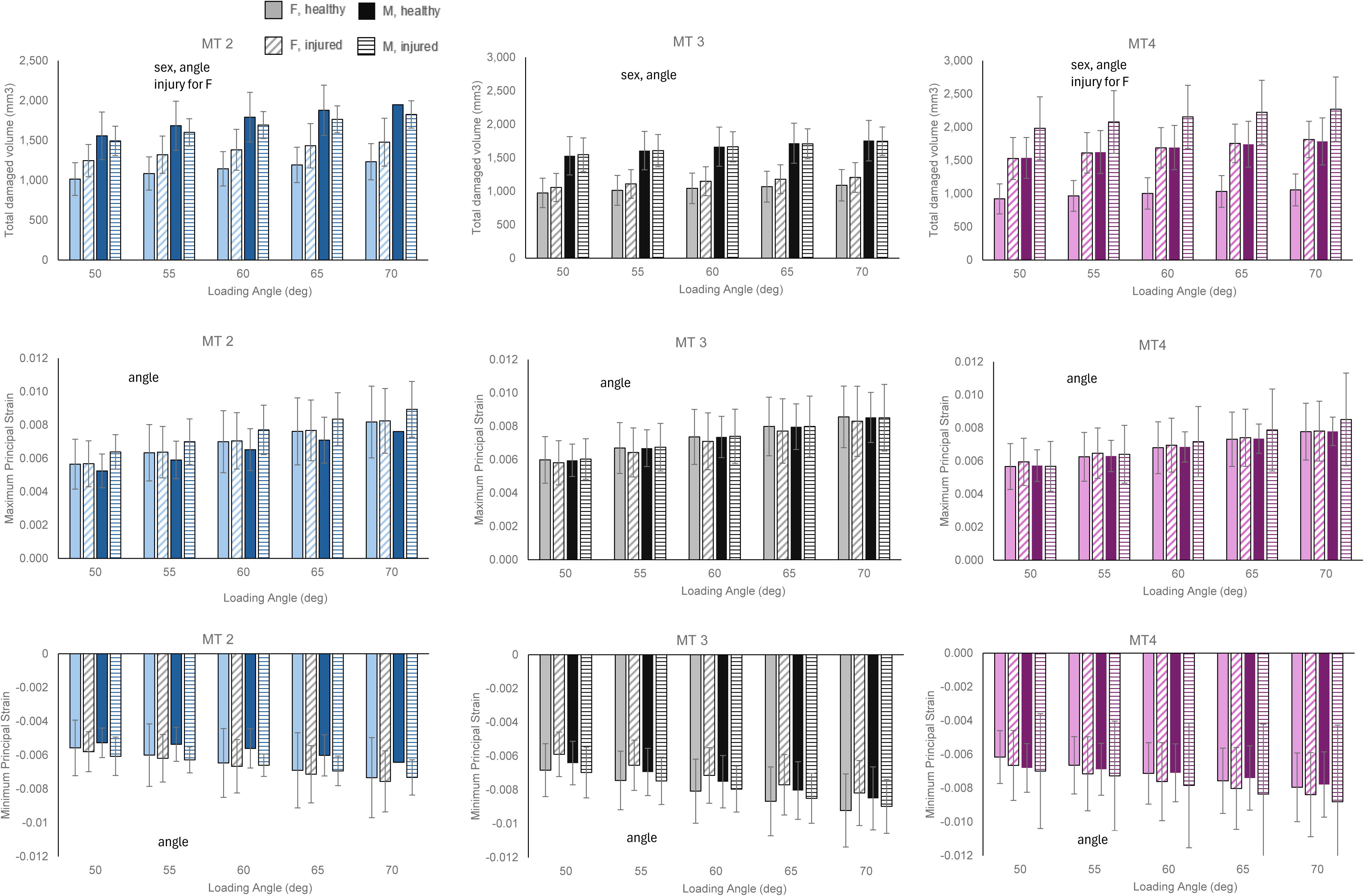
(Top row) Damaged volume, (middle) maximum principal strain, and (bottom) minimum principal strain for MT2 (left / blue), MT3 (middle / black) and MT4 (right / purple) across loading angles 50° - 70°. Within each cluster we show F healthy, F injured, M healthy and M injured, with error bars indicating SD. Significant effects of sex, angle, and injury are indicated within each plot.

## Discussion

The purpose of this study was to clarify interactions between metatarsal structure, mechanical behavior, trainning volume, and assocation with BSI history in runners with and without metatarsal BSI. We hypothesized that metatarsal structure would scale with body size but not training volume, and that this scaling would not be affected by injury status. This hypothesis was generally supported by our data. Bone volume, BMC, CSA, and strength indices all scaled with both height and weight but not training volume. However, BMD did not scale with body size. This is not surprising given that both BMC and BV scale similarly. While metatarsal BMD could in theory be an indicator of poor bone quality, we observed no differences in BMD values between participants or within subgroups.

Our hypothesis that injured runners would exhibit lower metatarsal bone quality and mechanical properties compared to healthy runners was only partly supported. Our sample consisted largely of female runners. Injured female runners had slightly larger metatarsals with similar or slightly greater strength indices compared to their healthy counterparts. This was particularly apparent in the mid-shaft of MT4. At the same time, our FE analyses showed that injured female runners had similar principal strains, but higher damaged volumes than their healthy counterparts. This somewhat contradictory result might be explained by two things. First, although strength indices are important indicators of bone structure and quality, they are an average across the bone and not able to fully predict how an entire complex bone structure behaves under loading. In the tibia for example, we showed that FE models with multi-axial failure criteria better predicted fracture strength than regression models based on QCT variables including strength indices [26]. Second, damaged volume is likely related to overall bone size. We observed that injured female runners had slightly larger bones than their healthy counterparts, and that males had larger bones than females. Each of these subgroups also had greater damaged volume. Indeed, the observation that larger volume specimens of bone fail in fewer loading cycles than smaller specimens is the basis for using a measure of strained or damaged volume as a surrogate for fatigue failure life [13]. We hypothesize that the injured female group, which had higher training volume than the healthy group, had positive adaptations to their training that resulted in larger bones. Unfortunately, these adaptations were insufficient to withstand the demand placed on the bones by those runners. It is unclear from the present analysis which speicfic structural features within the metatarsals result in the observed behavior of increased strained volume without increased principal strains. It is also important to remember that the present analysis was of the uninjured metatarsals of runners with a metatarsal BSI, and there may be isolated structural deficits in the specific metatarsals that were injured. Future analyses of the injured bones themselves may provide more insights into the role of bone structure in metatarsal BSI.

Our relatively small group of male runners showed less consistent and sometimes opposite trends to our female runners. In general, the injured males had slightly smaller and weaker bones compared to their healthy counterparts, although these differences were not statistically significant in our sample. On average the injured male runners were nearly 6 kg lighter than the healthy runners, which may contribute to the observed differences. We only observed consistent (but still not significant) differences between injured and healthy males in the strained volume of MT4. Overall, our data do not point to a clear deficit in bone quality in injured male runners.

Our FE simulations applied a fixed percentage of bodyweight to the distal end of each metatarsal to simulate a mechanical test that was biomechanically relevant. The magnitude of the strains we observed were relatively high, which may be a reflection of our rigid constraint at the proximal metatarsal end. Our models do not consider how each metatarsal interacts with other structures within the foot. Indeed, load transmission within the structures of the feet, and the exact manner in which the metatarsals are loaded *in vivo* is not currently well understood. Peak compressive strains ranging from -1500 to -2300 micro-strain have been measured on the dorsal MT2 during barefoot walking using strain-gage staples [27]. These ranges increased to -5300 micro-strain during jogging [28], a value similar to our calculated compressive strains in the 50° loading condition. However, the forces applied to each specific metatarsal during running could be altered through changes in running biomechanics, footstrike mechanics, and potentially even footwear.[30]

We consistently observed that both bone strain and damaged volume increased with greater loading angle in our simulated mechanical tests. In theory, a person could modify the angle of the resultant force applied to the metatarsals through the activation of the plantar intrinsic muscles, along with extrinsic muscles that support the foot arch. A higher arch and more active muscles are expected to result in a more axial load (decreased loading angle), which would reduce bone strain and subsequent damage. Our data demonstrate that a 10 degree change in loading direction reduces metatarsal strain by 22%. The degree to which this could be achieved through training or by altering footstrike biomechanics or footwear is yet to be determined.

Within-human characteristics of both bone quality and mechanical behavior are influenced by multiple factors including history of physical activity, diet, and other factors. In this cohort, the average age of purposeful running was 15 and healthy male runners started earlier (at age 12). If one assumes mid-puberty (Tanner stage 3) at age 12 for women and 14 for men [29], then fewer female runners (5 out of 39) than male runners (11 of 14) started running prior to or during mid-puberty. Bone diameter and cross-sectional shape and area are influenced by physical activities during growth and adolescence, and bone accrual may be further influenced by adequate energy availability [30,31]. Furthermore, participation in multidirectional loading sports such as basketball and soccer during puberty has been suggested protect against future BSI [32,33]. While no data exist on the degree to which metatarsal structure in particular is influenced by early physical activity, the relatively late start date (relative to pubertal timing) would indicate that running was unlikely to have much influence over metatarsal geometry for most participants in this study. However, a more detailed analysis of physical activity and sports participation history could provide additional insights on ths topic. It is possible that the timing of running onset relative to puberty explains some of the male/female differences that we observed in our data or sex-differences in response to loading. These factors deserve further research to determine optimal strategies to optimize bone strength and quality during the critical time of puberty.

The few structural differences in the metatarsals of injured versus healthy runners suggest other factors may contribute to risk for BSI. Within our sample, the injured runners had significantly lower BMI and higher weekly distance than the healthy runners. Low BMI is a biological/physiologic risk factor associated with relative energy deficiency syndrome (REDs) [9]. REDs and the terminology of Female Athlete Triad [34,35] describe the influence of low energy availibility to hormones including estradiol, and both of these factors contribute to impaired bone health. Low energy availability is defined as the difference of energy intake to exercise energy expenditure, standardized to fat-free mass, and is more common in female than males, particuarly for endurance runners [36]. This can affect both the present bone structure, which we measured here with CT, as well as the ability of bone to adapt and repair damage. Our CT data do not show any consistent deficits in metatarsal bone structure. Our cohort of injured runners had lower BMI than their healthy counterparts, but the average is still above the BMI that would be considered a risk factor.

Not surprisingly, our injured cohort had significantly higher weekly distance than the healthy runners, and high training volume is a known risk factor for BSI. The higher weekly volume of training increases cumulative load to bone and may contribute to low energy availability state if the increase exercise energy expenditure is not addressed with increased energy intake. Many runners successfully maintain high training volumes without injury, indicating that they are either running in a manner that reduces metatarsal damage or are physiologically able to adapt to high training volumes more successfully than their injured counterparts. Bones structurally adapt to habitual loads through osteocyte sensation of mechanical strain [37]. These cells are effective in responding to mechanical stimuli but become desensitized over time[38]. Specifically, after 30 minutes of intense training, bones become desensitized to their stimuli and no longer maximize bone formation, but as a structure and material they continue to accumulate damage [4,39]. Our running history survey did not clarify how training volume is divided by sessions over a typical week or vary throughout the year. In theory, one could divide up weekly distance into more frequent, shorter bouts to maximize adaptation by preventing osteocyte desensitization. In fact, ultra-runner Camille Harron, who maintains very high weekly training volumes injury-free, made a claim that she has implemented this exact strategy with good success [40].

There are a few limitations to our study. First, because the majority of our participants were female runners, we had smaller number of male runners, which may explain our inability to detect differences between injured and healthy males. Second, we used FE modeling here to simulate a simple biomechanically relevant mechanical test that was scaled to body weight. Our simulations do not account for the loading distribution across the metatarsals, footstrike pattern, varying use of intrinsic foot muscles, and numerous other factors that affect the magnitude and direction of the forces applied to the metatarsals. We did not analyze the elements close to the constraints and applied loads to avoid artifact in the models, and therefore our findings are only relevant to the mostly-cortical diaphyses of each metatarsal. Our simulation data should be interpreted as an indicator of metatarsal structural behavior, but the specific values of strain and damaged volume do not necessarily represent what occurs during the stance phase of running. Furthermore, our measure of Tsai Wu damaged volume as an indicator of bone fatigue life has not been validated with mechanical testing in human metatarsals. Our study also has several strengths. Although our sample of recently injured runners was relatively small, we statistically powered our study by collecting a larger healthy comparison group. To our knowledge, this is the most comprehensive study reporting on metatarsal bone structure and mechanics that we know of. Our runners varied from recreational runners to competitive NCAA Division I collegiate athletes, making this cohort a good representation of runners in general. And finally, our unique approach to simulation provides information that can be used to generate strategies for BSI risk reduction through mechanisms of reducing metatarsal strain and subsequent damage.

In summary, we report here on metatarsal structure and mechanical behavior in male and female runners with and without a recent bone stress injury. We found that bone size and strength scales with body mass and height. We observed that injured female runners had slightly larger metatarsals with increased strength metrics than their healthy counterparts, which we believe reflects adaptation to higher training volumes. Despite their larger volume and higher bone mineral content, the metatarsals of injured female runners had similar mechanical strain and greater damaged volume compared to healthy runners, supporting the value of FE simulations to provide mechanically relevant information that is independent from QCT analyses. Our simulations demonstrate a significant and consistent effect of loading angle on all strain and damage metrics, indicating that reducing metatarsal bending through the activation of plantar intrinsic muscles or other mechanisms may represent a method for reducing risk of metatarsal BSI.

## Data Availability

All data produced in the present work are available upon reasonable request to the authors.

## Acknowledgement

This research was supported by a Midcareer grant from the Foundation for Physical Medicine and Rehabilitation (to AST) and by NICHD of the National Institutes of Health under award number R15HD104169 (to KLT, with supplement to BAY). The content is solely the responsibility of the authors and does not necessarily represent the official views of the National Institutes of Health. The results of this study are presented clearly, honestly, and without fabrication, falsification, or inappropriate data manipulation.

## Notes

### Competing Interest Statement

The authors have declared no competing interest.

### Author Declarations

The Ethics committee/IRB of Worcester Polytechnic Institute gave ethical approval for this work.

